# AutoClip: AI-Guided TEE Semantic Segmentation for TEER A Proof-of-Concept Study

**DOI:** 10.64898/2026.05.29.26354195

**Authors:** Mi Chen, Xia Li, Kairui Yang, Maurizio Taramasso

## Abstract

**Background:** Transcatheter edge-to-edge repair (TEER) is an established treatment for mitral regurgitation but remains highly dependent on operator experience and complex transesophageal echocardiography (TEE)-guided intraprocedural imaging. Artificial intelligence (AI)-based semantic segmentation may improve procedural reproducibility and intraprocedural guidance; however, no TEER-specific segmentation framework has been reported.

**Objectives:** To develop and evaluate AutoClip, a clinician-driven AI-guided TEE semantic segmentation model designed for simultaneous delineation of mitral valve anatomy and in-vivo TEER device components.

**Methods:** A retrospective proof-of-concept study was conducted using 987 intraprocedural TEE frames derived from 10 video clips in 3 patients undergoing MitraClip G4 implantation. Seven semantic labels, including mitral leaflets and device components, were manually annotated using ITK-SNAP. Following standardized preprocessing and region-of-interest extraction, an Attention U-Net architecture was trained frame-wise on bicommissural and corresponding X-plane TEE views. Model performance was assessed using mean intersection-over-union (IoU) and Dice coefficient on an independent test set.

**Results:** The Attention U-Net demonstrated improved sensitivity to small device structures compared with conventional U-Net architectures. Preliminary training performance achieved a mean IoU of approximately 0.93, while independent test performance reached a mean IoU of 0.46 across foreground classes. Qualitative assessment demonstrated feasible simultaneous segmentation of mitral leaflets, clip arms, grippers, and delivery shaft during TEER procedures.

**Conclusions:** AutoClip represents a proof-of-concept TEER-specific TEE semantic segmentation framework initiated through a clinician-oriented workflow without formal computer science expertise. Although preliminary accuracy remains modest due to limited sample size, this study establishes a reproducible pathway for future AI-assisted intraprocedural guidance systems and larger multicenter development efforts in structural heart interventions.

---

Transcatheter mitral edge-to-edge repair (TEER) is an established therapy for inoperable patients with degenerative mitral regurgitation and for those with functional mitral regurgitation associated with reduced left ventricular ejection fraction (LVEF)(1). Despite its growing evidence, TEER remains technically challenging concerning transesophageal echocardiography (TEE)-guided intraprocedural imaging, where difficulties stem from echocardiographer experience, variability in patient anatomy, seamless communication between operators and echocardiographers. Artificial intelligence (AI)-based solutions have the potential to mitigate these sources of variability and provide more predictable results across diverse clinical scenarios. Although echocardiography-based semantic segmentation has been applied to mitral valve anatomy(2), no TEER-specific segmentation model has been reported. Here, we introduce **AutoClip**, an AI-guided TEE semantic segmentation model designed to delineate both mitral valve anatomy and in-vivo TEER devices. Notably, we present a step-by-step, clinician-oriented framework for initiating computer vision-based modeling, demonstrating feasibility without a computer science background, followed PRIME checklist(3):

We conducted a retrospective proof-of-concept study including ten intraprocedural transesophageal echocardiography (TEE) video clips (987 frames) from three patients who underwent TEER with MitraClip G4 devices (Abbott). All patients provided written informed consent, and the study was approved by the Kanton Ethics Committee (2025-00443). Only bicommissural views and corresponding X-plane images, in which the MitraClip was visualized capturing the leaflets, were included.

## Step 1: Data Standardization

Manual segmentation was performed using ITK-SNAP software (Version 4.0.1). Seven labels (clear background, anterior leaflet, posterior leaflet, anterior gripper, posterior gripper, clip arm, and shaft) were used to delineate mitral valve anatomy and MitraClip profiles, with all annotations performed by two experienced TEER physicians. DICOM TEE videos were paired to corresponding framewise NifTI masks by matching image tags (renaming non-“.dcm” DICOMs where needed). Pixel data were read as grayscale; if RGB stacks were present, the first channel was retained. Videos were organized as tensors of shape (T, H, W). Corresponding masks were loaded from NIfTI and axis-aligned to the video (transposed from (W, H, T) to (T, H, W) when required). To focus on the tooling/anatomic region, a fixed region of interest was applied (height 35–94% and width 55–98% of the original frame), after which frames and masks were resized to 256×448. Image intensities were scaled to [0, 1]; masks were resampled with nearest-neighbor interpolation to preserve labels. Class imbalance was quantified and addressed by computing median-frequency class weights across the seven labels. Debug overlays were periodically saved to verify image–mask alignment and preprocessing quality.

## Step 2: Model Selection

We briefly evaluated SAM2 but excluded it due to the need for extensive fine-tuning and per-frame prompting, which was impractical for this feasibility study. A standard U-Net was trained and achieved reasonable anatomy segmentation but struggled with small, low-contrast device parts. We therefore adopted an Attention U-Net, which applies attention gates to each skip connection to suppress irrelevant background and enhance critical leaflet–device interfaces. This architecture improved sensitivity to small structures while remaining lightweight and feasible for our limited dataset.

## Step 3: Model Assessment

Models were trained frame–wise on Apple Silicon (MPS) with Adam (lr□=□1×10□□) for 40 epochs (batch size□=□2; per–video variable lengths handled by zero–padding in the time dimension). Inputs were 1×256×448; outputs comprised seven classes. The optimization used unweighted cross–entropy (background class□=□0), and performance was tracked each iteration with a macro mean IoU computed over foreground classes 1–6 (background excluded). Checkpointing enabled resume–training, and qualitative assessment was performed by saving triptychs (input/prediction/ground truth) for the first three batches of each epoch; loss/IoU curves and a compiled DOCX report were generated automatically. Although median–frequency class weights were computed from the dataset to address imbalance, they were not applied in this feasibility run. With the limited dataset and no held–out split, results are reported as preliminary training–set performance (mean IoU ≈–0.93), consistent with our letter’s focus on process initiation rather than mature accuracy.

## Step 4: Model Evaluation

The trained Attention U-Net was evaluated on an independent test set using mean intersection-over-union (IoU) and mean Dice coefficient, calculated across six foreground classes (background excluded). Predictions were generated frame-by-frame and compared to manually annotated ground truth masks. Representative frames (start, mid, end) were visualized to assess qualitative performance. The model achieved a mean IoU of 0.46, reflecting preliminary accuracy given the limited dataset and the study’s focus on demonstrating feasibility of model initiation rather than optimized performance.

In summary, this proof-of-concept study demonstrates the feasibility of initiating a TEER-specific TEE semantic segmentation model from a clinician-driven workflow without computer science expertise. While preliminary performance was modest due to the limited dataset, the AutoClip framework establishes a reproducible pathway for future development, enabling expansion to larger, more diverse datasets and potential integration into intraprocedural guidance systems (Figure 1).

**Figure 1.**
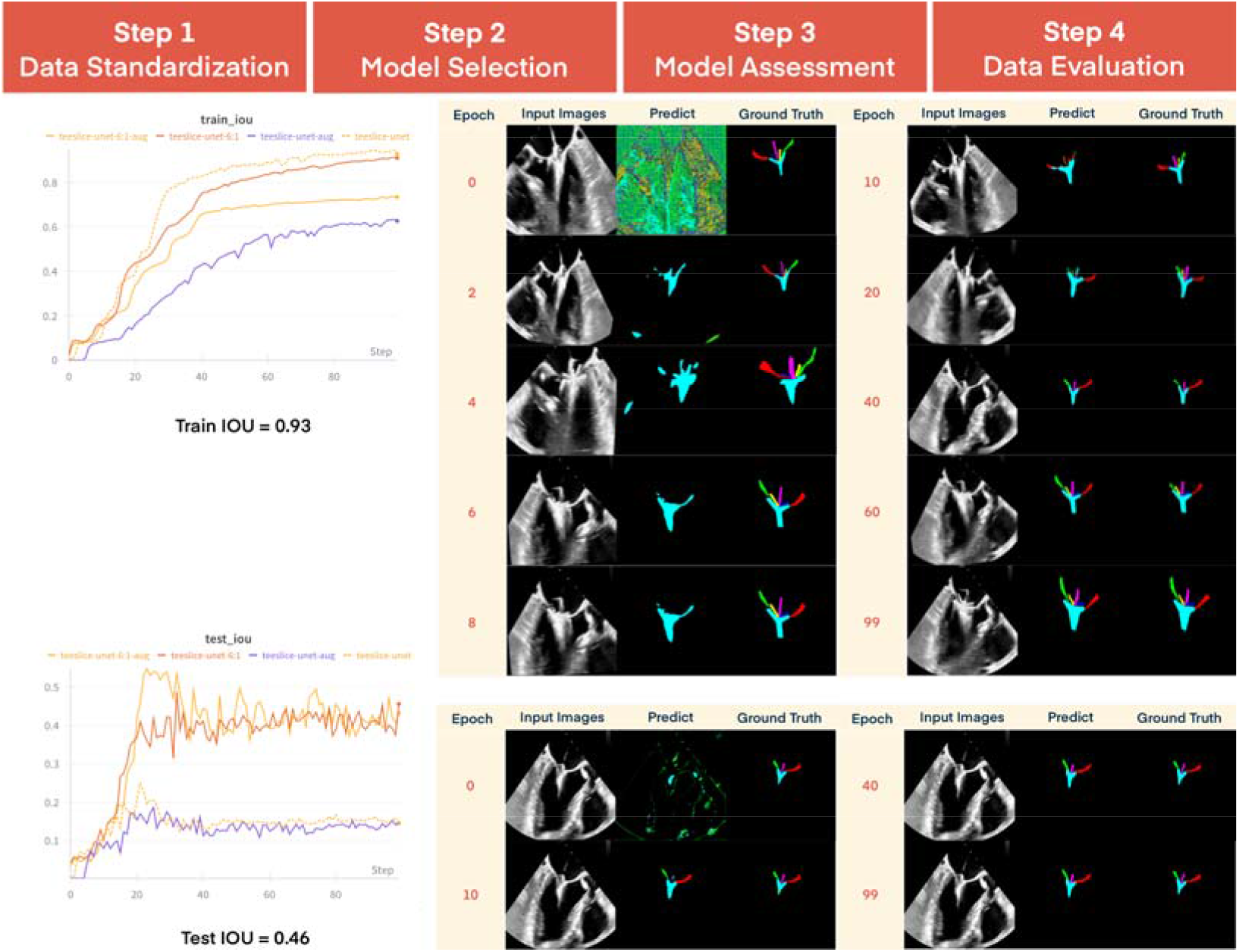
Workflow and Initial Results of AutoClip Model. The figure illustrates the workflow of the AutoClip semantic segmentation model, including data standardization, model selection, model assessment, and model evaluation. The lower left panel shows the training performance with a mean intersection-over-union (IoU) of 0.93 and a test IoU of 0.46. The right panel demonstrates the evolution of predicted segmentation masks over 0–100 epochs compared with the corresponding ground truth, while the lower right panel displays representative test image predictions across the training process.

## Data Availability

All data produced in the present study are available upon reasonable request to the authors

